# Do self-management interventions improve self-efficacy and quality of life in stroke survivors? A literarture review

**DOI:** 10.1101/2022.01.23.22269724

**Authors:** Yurike Septianingrum, Siti Nurjanah, Moses Glorino Rumambo Pandin

## Abstract

**Objective:** The aim of this review was to assess the effectiveness of self-management interventions on the self-efficacy and quality of life of stroke survivors compared to usual care.

**Method:** Article searches were performed using the same keywords in the Pubmed, CINAHL, Scopus and Science Direct databases published from January 2020 to October 16, 2021. A total of 8 articles in randomized controlled trials were identified in this study.

**Findings:** Eight studies were taken, of which six discussed the effectiveness of self-management interventions on self-efficacy and two literatures measured the quality of life of stroke patients. Almost all participatory studies reported an increase in self-efficacy and quality of life after receiving the self-management intervention. The heterogeneity in this study is reported regarding the form of intervention, duration of implementation, instruments used, and outcomes

**Conclusions:** There are various self-management interventions, which can be in the form of education, support, coaching, or empowerment. Self-management interventions are reported to improve self-efficacy and quality of life of stroke patients. Future studies are expected to measure the perceived burden of self and self-care in stroke patients

## INTRODUCTION

Stroke is one of the leading causes of death worldwide, the third leading cause of death, and one of the most expensive health problems in the world (Allen et al., 2010; Venketasubramanian et al., 2017). Ischemic stroke (53.7%) was the more common type, and hypertension (76.8.3%) was the main risk factor. First stroke 3 episodes (Tini et al., 2020). Long-term disability resulting from stroke poses a substantial health care burden (Sit et al., 2016). The burden of health, economic and social costs, has increased for stroke patients, their families, and the national health care system (Y. Chen et al., 2021).

One of the interventions that can be applied to stroke patients is self-management training as a promote and preventive effort to prevent recurrent strokes (Nott et al., 2021; Whitehead, 2018; Wolf et al., 2017). Prevention of recurrent stroke can be done through self-management interventions that involve patients in the process of changing their health behavior. Self-management refers to an individual’s ability to manage the symptoms, medication, physical and psychosocial consequences and lifestyle changes inherent in living with a chronic condition (Sakakibara et al., 2021).

Several reviews have shown the effectiveness of improving self-management in patients with chronic diseases (Hanlon et al., 2017; Ko et al., 2018), while reviews that discuss the effectiveness of self-management in stroke patients are still limited. In a review conducted by Sakakibara et al (2021) described self-management interventions used to improve risk factor control in stroke patients (Sakakibara et al., 2021), whereas Pedersen et al (2020) conducted a systematic review to determine the efficacy of self-management interventions for people with stroke over the age of 65 in relation to their psychosocial conditions (Pedersen et al., 2020). From all the literature reviews, there was no literature review that describes the effectiveness of self-management on self-efficacy and quality of life of stroke survivors.

Self-management provides clients with knowledge and skills that increase confidence, self-efficacy, and motivation to actively manage their ongoing recovery and rehabilitation (Nott et al., 2021). Self-efficacy and social cognition theory form the basis of many self-management programs and the link between self-efficacy and quality of life for stroke patients. (Jones & Riazi, 2011). Self-management is a treatment approach that allows individuals to solve problems as they arise, practice new health behaviors, and gain emotional stability (Sajatovic et al., 2018). Components of a self-management intervention after a stroke may include problem solving, goal setting, decision making, self-monitoring, coping with the condition, or interventions that maintain or improve physical and psychological functioning. (Whitehead, 2018). The purpose of this literature review was to assess the effectiveness of self-management interventions on self-efficacy and quality of life of stroke survivors.

## METHOD

### Eligibility criteria

PICOS criteria (Population, Intervention, Comparison, Outcome, Study type) were used to develop eligibility criteria for study inclusion and exclusion in a randomized controlled trial review. (Eriksen & Frandsen, 2018). The criteria are:

P (Population): stroke patient

I (intervention): self-management

C (Comparison): usual care

O: self-efficacy and quality of life

S: randomized controlled trials

### Search strategy

The literature search was carried out using an electronic database, limited from January 1, 2020 to October 16, 2021. The databases used to search for literature were Scopus, PubMed, CINAHL, and Science Direct. Search is limited to the use of English. The keywords used in the literature search were stroke” OR “cerebrovascular accident” AND “self-management” OR “stroke self-management” AND “self-efficacy” AND “quality of life” AND “randomized controlled trial” OR “RCT”. The same keywords were used in the literature search in each database. Boolean operators are used to combine keywords and index terms, and search results are refined using filters depending on each database.

### Study selection

All citations retrieved during the search process are exported to Mendeley, then the collected citations are filtered to remove duplicates. Notes are then filtered through titles and abstracts to exclude review articles and adjustments to the criteria. The article’s feasibility study is carried out by reviewing full-text articles. Articles deemed appropriate by the reviewers were used in this literature review. The article selection process and results are presented in the PRISMA diagram in (figure 1).

**Figure 1.**
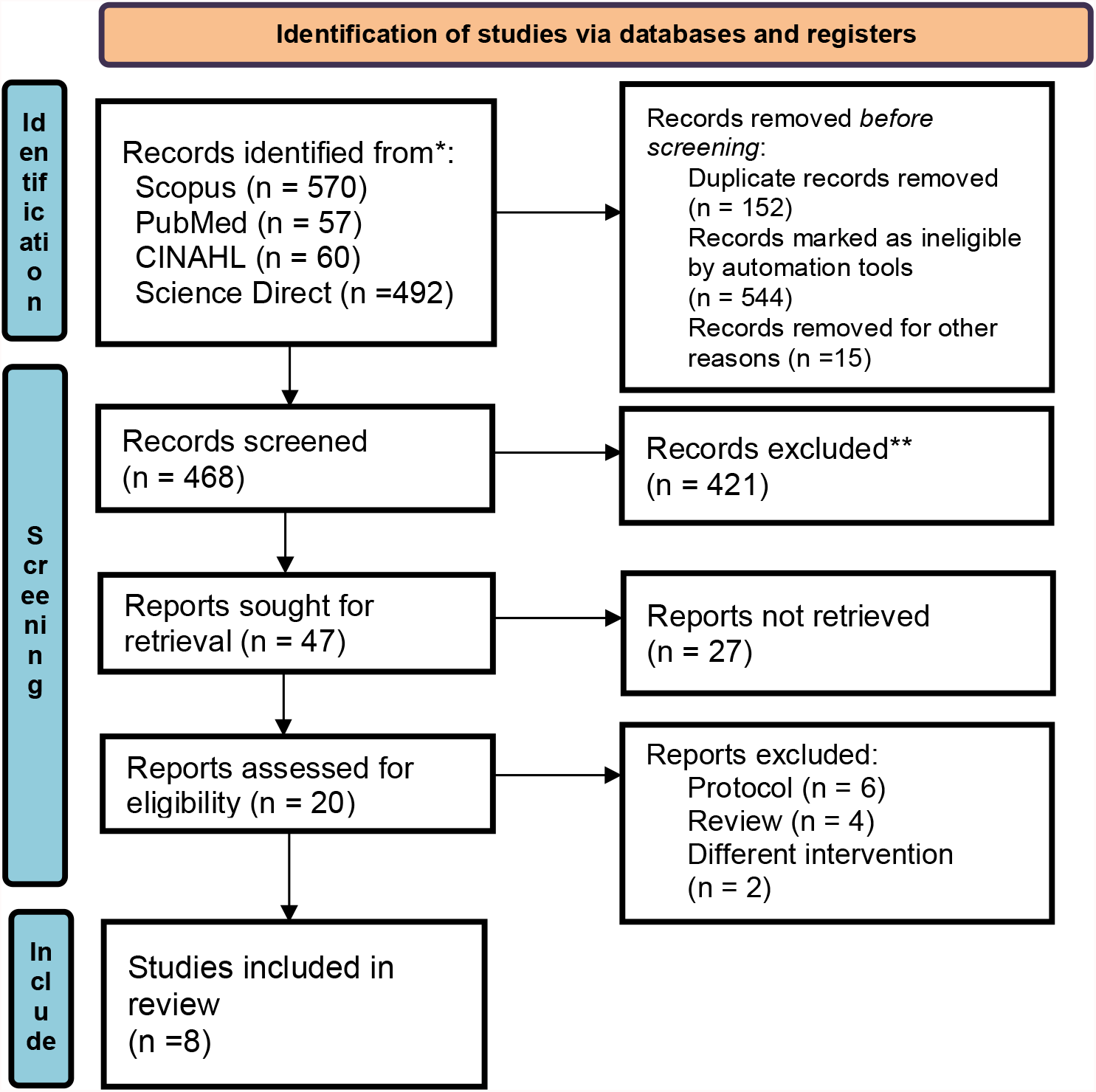
PRISMA Flowchart of Literature Search and Screening Process

## RESULT

A search through four databases yielded 1966 citations, which were then filtered to exclude duplicates, filtered focusing on stroke cases, and the population in non-children, yielding 468 records. A total of 47 records were obtained based on PICOS, namely P: stroke patients, I: self-management, C: usual care, O: self-efficacy and quality of life, S: randomized controlled trials. A total of 20 full-text articles were assessed for eligibility, and only 8 RCT articles matched eligibility in the quantitative study.

**Table 1.**
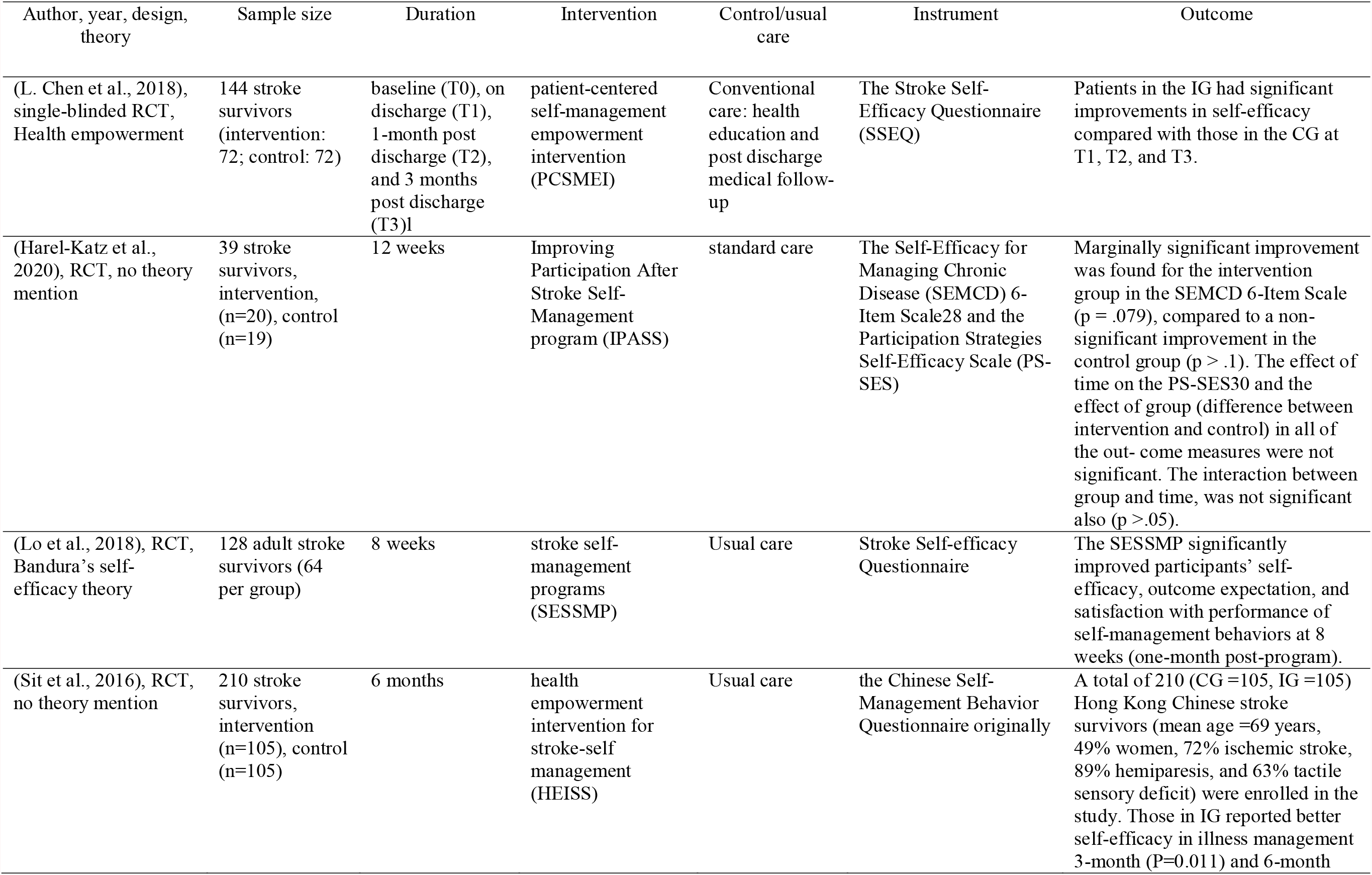
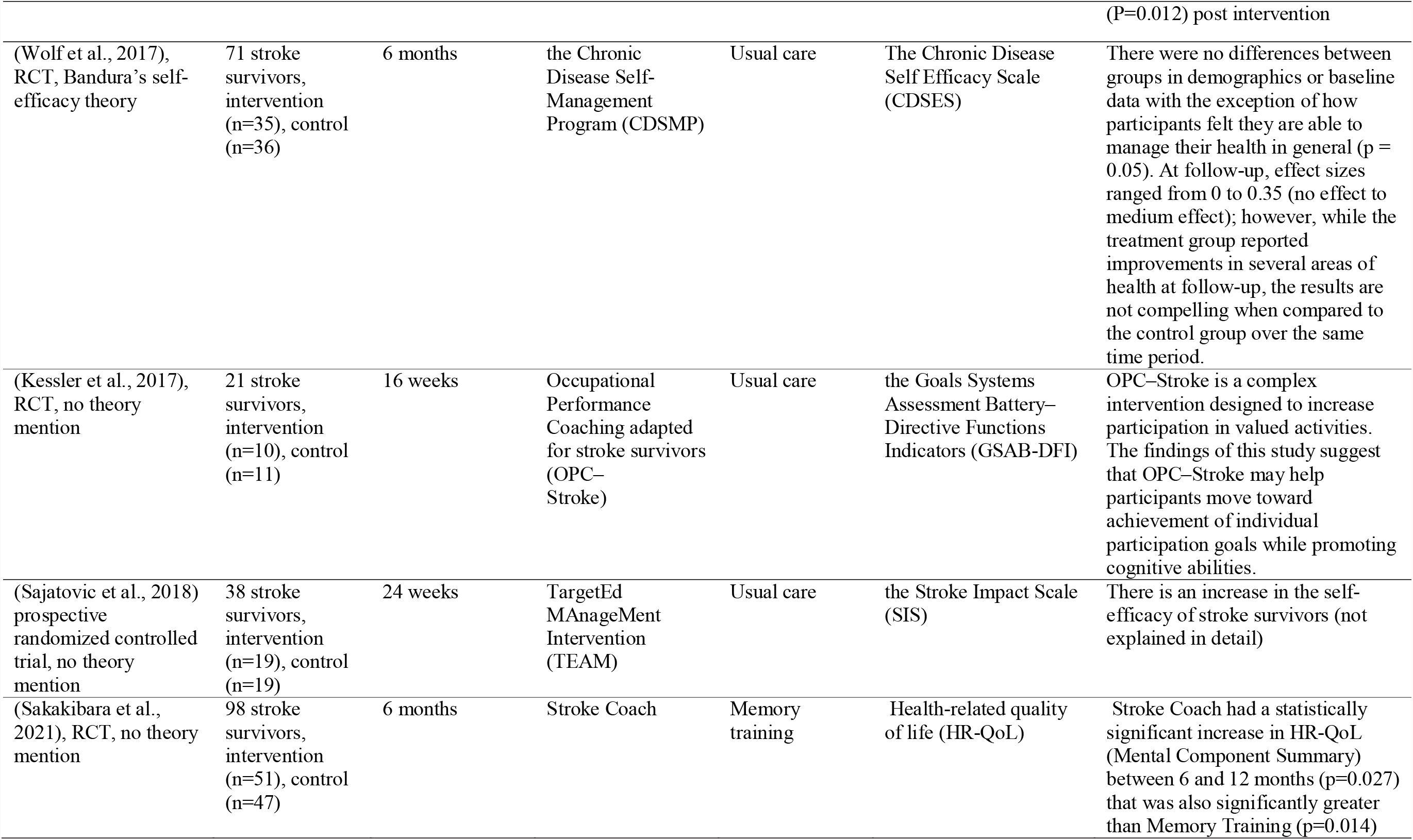
Characteristics of the literature included in this review (n=7)

## DISCUSSION

This study focuses on stroke patients, where stroke patients experience a disability at risk of death (Allen et al., 2010). About 15 million people suffer a stroke every year, most of them are ischemic due to modifiable risk factors (Ortiz-Fernández et al., 2019). Stroke is the leading cause of long-term disability worldwide. Several studies have shown that stroke has a negative effect on participation, which is manifested by difficulty returning to meaningful daily activities several months and even years after the stroke. (Harel-Katz et al., 2020)..

A total of 5 of the 7 literatures discussed in this literature review show that self-management interventions have a positive effect on self-efficacy (Harel-Katz et al., 2020; Lin et al., 2020; Lo et al., 2018; Sit et al., 2016; Wolf et al., 2017). This shows that self-management plays an important role in increasing self-efficacy. So far, self-management interventions are identical to health education with the Pender’s health promotion model approach, and increasing self-efficacy with Bandura’s self-efficacy theory approach.

Several studies show the effectiveness of increasing self-management on self-efficacy (Lo et al., 2018; Nott et al., 2021) and quality of life of stroke survivors (Ortiz-Fernández et al., 2019; Sajatovic et al., 2018; Whitehead, 2018). Self-efficacy is defined as the belief and confidence that individuals feel in their ability to perform a particular task or action and increased self-efficacy is a desired outcome of a self-management program (Nott et al., 2021). Several articles used Albert Bandura’s concept of self-efficacy (Lo et al., 2018; Wolf et al., 2017) According to Bandura (Smith & Liehr, 2018), There are four main sources of self-efficacy: direct mastery experiences, vicarious experiences, verbal persuasion, and physiological states. Self-management of stroke patients draws on the work of several authors and includes the following concepts: a self-management approach provides clients with knowledge and skills that enhance self-confidence, self-efficacy, and motivation to actively manage their ongoing recovery and rehabilitation (Nott et al., 2021).

Almost all studies discussed in this literature review state that self-management interventions can increase self-efficacy in stroke survivors. A premise of self -management is that individuals who have a greater expectation that they are capable of performing a behavior to produce a given outcome are seen as having greater self-efficacy (Fryer et al., 2016). Self-management interventions for people after stroke that aim to increase individuals’ abilities to solve problems, make decisions, and construct action plans for specific functional targets, could help prevent some of the difficulties that people with stroke face when discharged from rehabilitative health care (Jones et al., 2016). The use of instruments to measure self-efficacy also varies, but only one article uses the Stroke Self-efficacy Questionnaire instrument that specifically measures self-efficacy in stroke survivors.

About a third of stroke survivors experience a mood disorder, most commonly depression, anxiety, and these conditions are the most stressful for families. Overall, these factors affect the quality of life (QoL) (Whitehead, 2018). Self-management is a treatment approach that allows individuals to solve problems as they arise, practice new health behaviors, and gain emotional stability (Sajatovic et al., 2018). Quality of life can be improved by self-management interventions that accomplish more than a single domain of change. The success of these interventions depends on participation levels, impairment of participant, health services’ use, health behavior, costs, participant’s satisfaction, and associated adverse events during the intervention period. However, these interventions are often difficult, time-consuming, and human resources intensive (Ortiz-Fernández et al., 2019). In the control group, almost all of them provided the usual care intervention, only two articles compared it with other interventions, where the self-management intervention was superior to other interventions, this became an added value to the self-management intervention and could be applied to stroke patients to reduce the risk of stroke improve their quality of life.

## CONCLUSION

This literature study discusses the effectiveness of self-management interventions in stroke patients with several parameters, but the similarities throughout the literature refer to self-efficacy. Self-management interventions are interventions that can be given to patients when the patient comes home from the hospital or when the patient is already at home. Stroke self-management led by nurses can increase self-confidence which makes them willing and able to play an active role in managing their own health and the effect of exercise on events that affect their lives during the stroke rehabilitation journey. It is hoped that the development of self-management coaching interventions can be integrated through a different theoretical approach, namely Meleis’s Transitional Care, which has not yet been studied. This allows for optimization of care while at home by increasing patient readiness starting from the time the patient is in hospital.

## LIMITATION

This study has several limitations, namely some articles do not mention the theory or framework that underlies the study, some articles also do not clearly explain the research instruments used and secondary outcomes.

## IMPLICATION OF FINDINGS ON NURSING PRACTICE

This literature review is expected to be used as input for nursing science, especially medical surgical nursing in determining appropriate interventions for stroke patients. Selection of the right intervention can help in the rehabilitation process of stroke patients, especially in meeting their basic needs so as to improve the quality of life of stroke patients.

## Data Availability

All data produced in the present study are available upon reasonable request to the authors

## ACKNOWLEDGEMENT

The authors thank to the Faculty of Nursing Universitas Airlangga for the facilities in this study.

